# Exploring Novel Kinetics of Automated H_2_O_2_ Nebulization: A Breakthrough in SARS-CoV-2 Elimination

**DOI:** 10.1101/2025.06.04.25327212

**Authors:** Jennifer Solano-Parada, José Antonio Sánchez-Martínez, Beatriz Carolina Gómez-Hernández, Margarita Barriga, Pilar García-Velasco, Alberto Fernández, Alberto Cornet-Gómez, Antonio Osuna Carrillo de Albornoz, Rene Fabregas, Concepción Morales-García

## Abstract

Although hydrogen peroxide (H_2_O_2_) nebulization has shown promise for reducing SARS-CoV-2 loads in healthcare settings, its precise kinetics and real-world efficacy remain incompletely understood. In this prospective environmental sampling study, we collected air and surface samples from COVID-19 patient rooms before and after automated H_2_O_2_ nebulization, quantifying viral RNA by RT-qPCR and viral antigens by indirect ELISA, while assessing infectivity via Vero E6 cell cultures. A piecewise exponential model characterized the kinetics of viral load reduction, capturing both initial delays and subsequent decay phases. Results revealed a marked decrease in RT-qPCR positivity rates, higher cycle threshold values indicative of lower viral loads, and substantially reduced cytopathic effects, suggesting that residual viral RNA was largely non-viable. These findings underscore the non-linear nature of H_2_O_2_-mediated decontamination and the influence of environmental variables such as airflow, humidity, and surface composition. By integrating molecular diagnostics, infectivity assays, and mathematical modeling, our study offers a comprehensive framework for refining decontamination protocols. Future investigations should explore larger, multi-institutional cohorts and evaluate the applicability of these insights to emerging viral threats in diverse clinical environments.

**Importance:** This study represents a significant advance in environmental decontamination by demonstrating that automated hydrogen peroxide (H_2_O_2_) nebulization markedly reduces SARS-CoV-2 contamination in hospital settings. By integrating rigorous molecular diagnostics with infectivity assays and sophisticated kinetic modeling, the research delineates the non-linear decay of viral loads, providing robust evidence that residual viral RNA post-treatment is largely non-infectious. Such insights are invaluable for refining disinfection protocols, ensuring patient and healthcare worker safety, and mitigating nosocomial transmission. Incorporating advanced machine learning techniques further improves the predictive accuracy of decontamination outcomes. Ultimately, this work lays a strong foundation for future studies to optimize and personalize disinfection strategies across diverse clinical environments, thereby bolstering public health efforts to combat current and emerging infectious diseases.

## 1 Introduction

The ongoing incidence of COVID-19 outbreaks within healthcare settings emphasizes the importance of implementing comprehensive disinfection protocols to reduce viral transmission effectively. Although respiratory droplets serve as the predominant mechanism for the spread of SARS-CoV-2, research indicates that contaminated surfaces and airborne particles significantly contribute to nosocomial transmission^1^. The sustained presence of the virus on hospital surfaces renders them potential reservoirs of infection, posing an ongoing risk to both patients and healthcare professionals^2^. Notably, the lipid envelope of SARS-CoV-2, which is crucial for its infectivity, is susceptible to the effects of oxidizing agents and surfactants^3^. Therefore, it is imperative that rigorous environmental cleaning and disinfection protocols are established as auxiliary measures in conjunction with droplet and contact precautions, forming an essential multilayered strategy for infection prevention within healthcare settings settings^4^.

Nebulization of hydrogen peroxide (H_2_O_2_) has emerged as a particularly compelling decontamination methodology for healthcare environments, warranting significant attention^5,6,7^. Its attractiveness is derived from its broad-spectrum antimicrobial activity, established efficacy against enveloped viruses, inherent practicality for deployment in enclosed settings, and ability to attack surfaces and airborne particles^8^. At the biochemical level, hydrogen peroxide exerts its effects through the formation of hydroxyl free radicals (OH•), which are highly reactive oxidants that induce cellular stress and inflict damage on critical microbial components, specifically lipids, proteins, and nucleic acids^9^. Indeed, vaporized hydrogen peroxide is well-documented for its capacity to effectively inactivate a wide array of viral and bacterial pathogens within hospital settings, including areas of limited accessibility^10^. While previous investigations have substantiated the virucidal properties of vaporized hydrogen peroxide against numerous microorganisms, including coronaviruses^2,11,12,13^, a comprehensive evaluation of SARS-CoV-2 inactivation under authentic hospital conditions remains imperative^14^. Therefore, a rigorous assessment of (H_2_O_2_) nebulization efficacy across varying concentrations and exposure times is critically needed to refine disinfection protocols and maintain the most stringent safety standards in healthcare environments^15^.

In the context of environmental surveillance concerning SARS-CoV-2, reverse transcription quantitative PCR (RT-qPCR) is frequently employed as the primary method for detecting viral RNA^16^. However, the exclusive reliance on RT-qPCR for environmental assessment presents a notable limitation. Although it exhibits sensitivity in detecting viral RNA, RT-qPCR cannot differentiate between intact, infectious viruses and inactivated, non-infectious viral fragments^17,18,19^. Consequently, the data derived from RT-qPCR alone cannot definitively ascertain the risk of infection associated with environmental contamination, nor can it adequately validate the effectiveness of decontamination efforts in mitigating infectious viruses. Viral culture methodologies are imperative to address this critical deficiency and to attain a more comprehensive understanding of environmental risk. By inoculating environmental samples onto Vero E6 cell cultures, it becomes possible to evaluate the infectivity of SARS-CoV-2 directly^1^. Significantly, the combination of RT-qPCR and viral culture techniques affords a powerful and nuanced approach to the issue^20^. This integrated strategy facilitates a more thorough assessment of viral persistence and infectivity following environmental contamination^21^. RT-qPCR quantifies the quantity of viral RNA present, while viral culture directly evaluates the infectious potential of that RNA. This amalgamated methodology presents a significantly more robust evaluation of environmental viral contamination than RT-qPCR when utilized in isolation. It is essential for accurately determining the efficacy of decontamination methods, ensuring that reductions in measured RNA levels correlate with diminished environmental infectivity and a lower risk of transmission. Although clinical diagnostic assays such as Spike protein-based Enzyme-Linked Immunosorbent Assay (ELISA)^22,23,24,25,26^ are critical in patient care and in enhancing the understanding of disease propagation, the combined RT-qPCR and viral culture approach is uniquely vital for ensuring accurate and comprehensive assessments of environmental risk.

In this study, we hypothesize that an automated hydrogen peroxide (H_2_O_2_) nebulization system can significantly reduce SARS-CoV-2 loads in hospital environments. Our primary objective is to clarify the non-linear kinetics of viral inactivation by systematically integrating RT-qPCR, infectivity assays, and mathematical modeling. First, we establish a strong framework for detecting both viral RNA and proteins in air and surface samples, thus evaluating the extent of contamination. Second, we apply quantitative infectivity assays in Vero E6 cells to validate the presence of viable SARS-CoV-2 and assess its susceptibility to H_2_O_2_. Third, we use piecewise exponential models to capture the complexities of virucidal kinetics, including delay phases and accelerated decay. The subsequent sections provide details of our experimental setup and highlight the gradual reduction in viral RNA and antigen loads post-decontamination. We then present a comprehensive discussion, connecting our findings to existing literature on aerosol stability and disinfectant efficacy. Furthermore, we address methodological limitations and consider how future research might enhance H_2_O_2_ nebulization protocols. Lastly, we outline potential clinical applications and emphasize the broader implications for infection control strategies. This paper offers a rigorous, data-driven evaluation of automated H_2_O_2_ nebulization, providing evidence-based insights into its effectiveness in reducing SARS-CoV-2 transmission risks.

## 2 Results

### 2.1 Room Sampling and Initial Virus Detection

A comprehensive environmental sampling study was conducted within 18 confirmed COVID-19 patient rooms at a tertiary care hospital, resulting in the collection of a total of 72 samples. This included 36 air samples and 36 surface swabs, obtained before (b_*t*_) and following (a_*t*_) the application of a standardized H_2_O_2_ nebulization decontamination protocol. To evaluate the presence of viral RNA and viral protein, two distinct methodologies were employed: reverse transcription-quantitative polymerase chain reaction (RT-qPCR) for viral RNA detection and an indirect ELISA, explicitly targeting the SARS-CoV-2 S2 protein, for viral antigen detection (see Table S1-S2 in the Supplementary Material). Initial analysis utilizing RT-qPCR indicated that, before decontamination, 20 out of the 36 air samples (55.6%; CI 95%: 38.1-72.1%) tested positive, exhibiting cycle threshold (C_*t*_) values ranging from 23.48 to 36.39—complete data are available in the Supplementary Material Table S3. Concurrently, 16 of the 36 surface swab samples (44.4%; CI 95%: 27.9-62.0%) were positive for viral RNA, with C_*t*_ values spanning 24.79 to 33.46. Following the implementation of H_2_O_2_ nebulization, a significant reduction in the detection of viral RNA was documented. Only 8 out of the 36 air samples (22.2%; CI 95%: 10.1-40.1%) remained positive by RT-qPCR (p = 0.014, Fisher’s exact test), and these exhibited elevated Cycle Threshold values (C_*t*_), indicative of lower viral RNA loads. Similarly, the number of positive surface swabs decreased to 5 out of 36 (13.9%; CI 95%: 4.7-30.0%; p = 0.028), with all positive samples revealing significantly increased C_*t*_ values (>30). These preliminary findings, derived from RT-qPCR analysis, illustrate a substantial and statistically significant decrease in SARS-CoV-2 RNA detection after the application of the H_2_O_2_ nebulization protocol, indicating a marked reduction in viral contamination levels within the patient rooms. However, low levels of residual RNA, as evidenced by high C_*t*_ values, remained detectable in a small subset of samples. The ELISA data, which will be discussed subsequently, offers complementary insights regarding the presence of viral protein. The efficacy of the decontamination protocol, as reflected in the changes observed pre- and post-treatment (see **Figure 3**), underscores the system’s capability to reduce airborne and surface contamination.

### 2.2 Viral Propagation and ELISA-Based Detection

To quantitatively evaluate the presence of infectious SARS-CoV-2 and the effectiveness of decontamination, Vero E6 cells were employed as a susceptible host. A volume of 500 *μ*L from each sample was inoculated onto Vero E6 monolayers and incubated at 37°C with 5% CO_2_ for one hour to facilitate viral adsorption. After removing the inoculum and a wash with phosphate-buffered saline (PBS), fresh medium was introduced, and the cultures were incubated for 48 to 72 hours at 37°C in a humidified environment containing 5% CO_2_. The assessment of viral replication was conducted through microscopic observation of cytopathic effects (CPE), characterized by phenomena such as cell rounding, detachment, and syncytia formation. Before the decontamination (b_*t*_), 25 out of the 72 cultures (34.7%; CI 95%: 24.2-46.7%) demonstrated mild to moderate CPE, which strongly correlated with positive RT-qPCR results. An indirect ELISA utilizing polyclonal antibodies against the SARS-CoV-2 S2 subunit was employed to quantify the viral antigens in the culture supernatants. The pre-decontamination (b_*t*_) samples revealed optical density (OD) values significantly exceeding the established positivity threshold (p < 0.05), indicating a substantial presence of the virus. Following decontamination (a_*t*_), only 4 of the 72 cultures (5.6%; CI 95%: 1.5-13.5%) exhibited minimal CPE. This reflects a statistically significant reduction in the frequency of cultures that displayed CPE. Moreover, the indirect SARS-CoV-2 S2 ELISA indicated an approximately threefold reduction in mean OD values after decontamination, with all post-decontamination sample OD values falling below the pre-determined positivity threshold, as illustrated in **Figure 1a**. The notable decrease in both the observation of CPE (representing an 84% reduction in the number of positive cultures) and the ELISA-based detection of viral antigens underscores the robust efficacy of the H_2_O_2_ nebulization protocol in diminishing the presence of viable, infectious SARS-CoV-2 to levels that fall below the detection limit of this in vitro assay, thereby effectively mitigating the potential for viral transmission within the tested environment.

**Figure 1.**
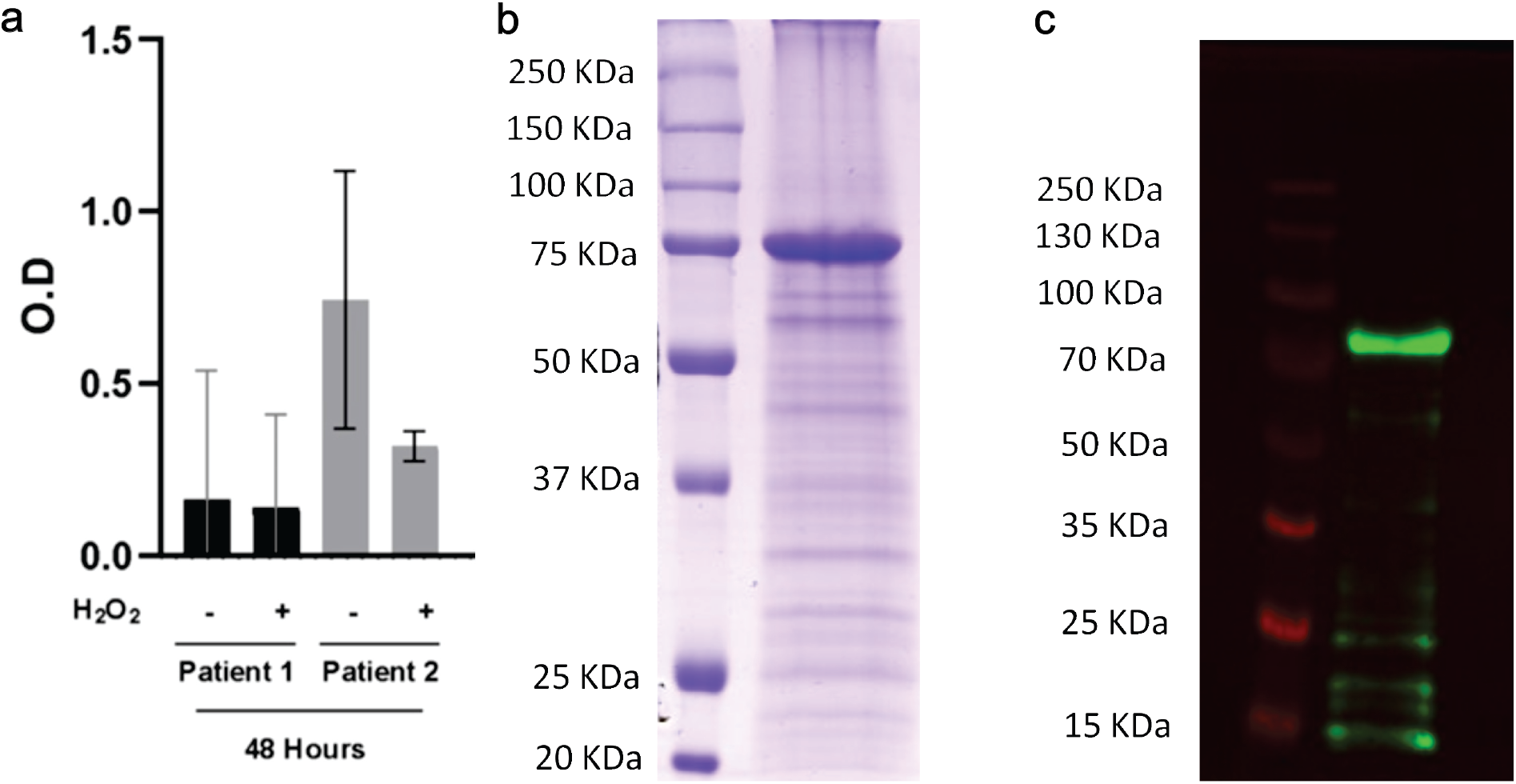
Evaluation of H_2_O_2_ Nebulization Efficacy and Verification of Recombinant SARS-CoV-2 S2 Protein Expression. **(a)** Air samples from two confirmed COVID-19 patient rooms were collected before (–) and after (+) treatment with a DuCFIT H_2_O_2_ nebulization system. Following a 1-hour incubation at 37°C to facilitate viral adsorption onto Vero E6 cells, the inoculum was replaced with MEM supplemented with 2% FBS and incubated for 48 hours. Viral S2 protein in the culture supernatants was quantified via indirect ELISA, with bar graphs representing mean optical density (OD) ± SD.**(b)** SDS-PAGE analysis of the recombinant SARS-CoV-2 S2-His6 protein. Lane 1 shows a prestained protein ladder, while Lane 2 displays the purified S2-His6 protein post-IPTG induction, with a prominent band at 75 kDa. **(c)** Western blot analysis confirms the expression of the S2-His6 protein. Proteins were transferred to a membrane and probed with an anti-His6 monoclonal antibody. Lane 1 contains the molecular weight marker; Lane 2 reveals a distinct band at the expected molecular weight (75 kDa), verifying the presence of the His6 tag.

### 2.3 RT-qPCR Analysis of N-Gene and S1/S2 Expression

Quantitative RT-PCR aimed at the SARS-CoV-2 N-gene was conducted utilizing the Genestore Detection Expert 1S kit. Following the protocols provided by the manufacturer, samples that exhibited a cycle threshold (C_*t*_) below 38 were classified as positive, while those displaying no amplification were assigned a C_*t*_ value of 0. Of the 72 environmental specimens collected (including 36 air samples and 36 surface swabs, as detailed in Section 3), 50% yielded detectable viral RNA, with C_*t*_ values ranging from 23.48 to 36.39 at baseline. Following the application of H_2_O_2_ nebulization, 58.3% of the previously positive samples transitioned to undetectable levels, while the remaining samples demonstrated statistically significant increases in C_*t*_ (p < 0.001, paired t-test), indicating a substantial reduction in viral RNA load. These findings align with the initial environmental sampling results and corroborate the significant decontamination efficacy documented by van Doremalen et al.^2^ and Chin et al.^3^. Furthermore, the consistently strong amplification of internal controls (mean *C*_*t*_ = 26.2 *±* 2.5) affirms both the integrity of the RNA extraction process and the minimal presence of PCR inhibitors, thereby enhancing the reliability of the RT-qPCR results.

SDS-PAGE of the induced cultures revealed prominent bands of approximately 75 kDa (**Fig. 1b**), which align with the predicted sizes of the His-tagged S1 and S2 subunits. Western blot analysis using an anti-His-tag antibody confirmed the expression of the recombinant proteins (**Fig. 1c**). Notably, the S1 and S2 subunits exhibited very similar electrophoretic mobilities, a finding that corroborates the observations of and underscores the necessity for additional techniques such as mass spectrometry for unequivocal subunit identification^24^.

### 2.4 Modeling Decontamination H_2_O_2_ Nebulization Kinetics

To rigorously dissect the complex temporal kinetics of hydrogen peroxide (H_2_O_2_) nebulization-mediated decontamination, a piecewise exponential model was deployed, explicitly accounting for the inherently non-linear nature of virucidal processes *(detailed theoretical formalism in Supplementary Document)*. This analytical framework, partitioning the decontamination process into distinct kinetic phases, is particularly well-suited to resolve the initial pre-decontamination stabilization period—potentially reflecting phenomena such as natural aerosol settling or nebulizer initiation dynamics—from the ensuing phase of exponential viral decay that signifies effective H_2_O_2_ virucidal action. Utilizing environmental SARS-CoV-2 load data –of Patient 2–summarized in **Table S1** of the *Supplementary Document*, we applied this piecewise model to parameterize the observed reduction in viral RNA following H_2_O_2_ nebulization. **Figure 2** graphically juxtaposes three distinct kinetic regimes—*rapid-onset with progressive deceleration* (**Fig. 2a**), *uniform reduction* (**Fig. 2b**), and *delayed onset with accelerated decay* (**Fig. 2c**)—each meticulously fitted to empirical RT-qPCR measurements (purple markers) and their experimental uncertainty (lavender uncertainty bands). For each regime, two primary quantitative parameters were derived through Non-Linear Least Squares fitting (NL-Fit): the virucidal rate constant (*k*, h^−1^), indexing decontamination efficiency post-delay, and the delay time (*τ*, hours), quantifying the pre-decay lag duration. Linear Least Squares fits (LS-Fit, dashed purple lines) were included for visual comparison of overall linear trends. To further validate model robustness and explore synthetic data generation, we employed a Generative Adversarial Network (GAN)^27^ trained on empirical data. Resultant GAN-generated synthetic data points (small violet markers, **Figure 2**) effectively interpolate between experimental measurements, providing a visually and statistically coherent representation of potential decontamination trajectories within each kinetic regime.

**Figure 2.**
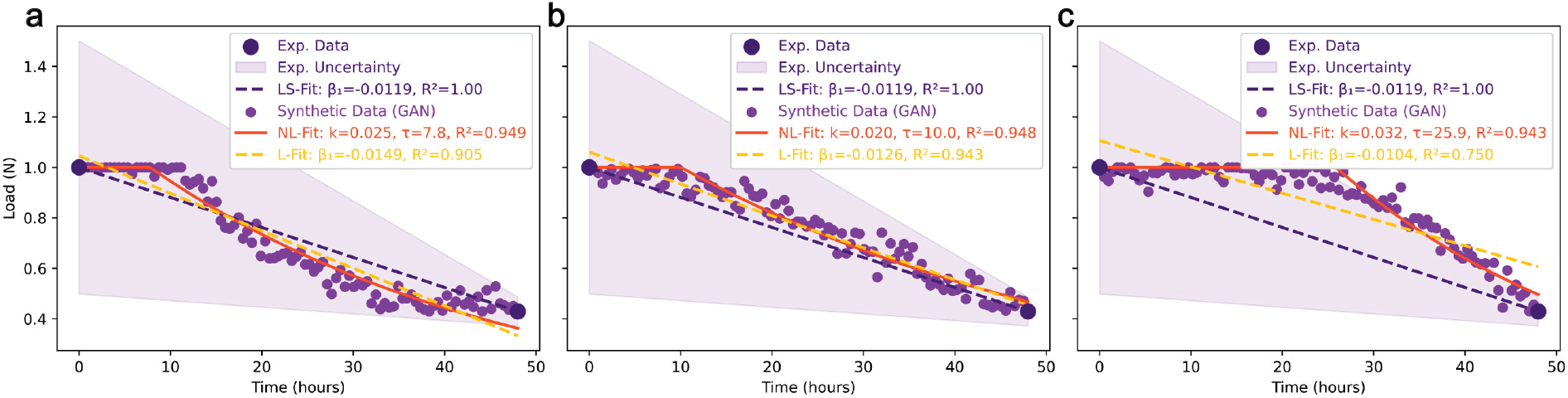
Comparison of three distinct kinetic regimes. Rapid-onset with progressive deceleration **(a)**, uniform reduction **(b)**, and delayed onset with accelerated decay **(c)**—applied to experimental data from Patient 2 (see Table S1, Supplementary). Purple markers and shaded uncertainty bands depict RT-qPCR measurements of SARS-CoV-2 load, with solid lines representing Non-Linear Least Squares (NL-Fit) and dashed lines indicating Linear Least Squares (LS-Fit). Orange markers denote GAN-generated synthetic data, illustrating the model’s capacity to interpolate effectively across the observed viral decay trajectories.

In the *rapid-onset with progressive deceleration* scenario (**Fig. 2a**), the piecewise exponential model –NL-Fit– yielded a rate constant of *k* ~ 0.025 h^−1^ and a delay time of *τ* ~ 7.8 h, achieving an *R*^2^ ~ 0.95 (Linear Least Squares Fit (L-Fit) *R*^2^ ~ 0.91). This parameter set reflects an initially swift decline in viral load that gradually tapers off, underscoring the importance of early-phase virucidal activity. By contrast, the *uniform reduction* scenario (**Fig. 2b**) produced *k* ~ 0.020 h^−1^, *τ* ~ 10.0 h, and an *R*^2^ ~ 0.95 (L-Fit *R*^2^ ~ 0.94), indicating a steady, nearly linear decay of viral RNA over the treatment period. Finally, the *delayed onset with accelerated decay* scenario (**Fig. 2c**), showed a markedly extended delay time (*τ* ~ 25.9 h) but a higher rate constant (*k* ~ 0.032 h^−1^), yielding an *R*^2^ ~ 0.94 (L-Fit *R*^2^ ~ 0.75). This profile is characterized by a protracted lag before H_2_O_2_ exerts its full virucidal effect, followed by an accelerated reduction in viral load. In all three cases, the piecewise exponential fits (NL-Fit) outperformed their corresponding Linear Least Squares approximations, as evidenced by systematically higher *R*^2^ values. Notably, in the last scenario (**Fig. 2c**), the linear fit achieved an *R*^2^ ~ 0.75, while the exponential fit showed a higher *R*^2^ ~ 0.94. These findings emphasize the non-linear nature of H_2_O_2_-mediated decontamination kinetics and underscore the value of including a delay parameter (*τ*) and a curvature factor (*c*) in modeling virucidal efficacy. Taken together, these scenarios – *rapid-onset with progressive deceleration, uniform reduction*, and *delayed onset with accelerated decay* – offer a robust quantitative framework for understanding the variability observed in experimental SARS-CoV-2 load reduction.

Furthermore, **Figure 2** delineates the viral decay kinetics by comparing the linear slopes (*β*) estimated via the intelligent GAN algorithm (L-Fit) against the constant experimental LS-Fit benchmark of −0.0119. In the rapid-onset deceleration scenario (**Fig. 2a**), the L-Fit yields a *β*_1_ of −0.0149, the steepest in absolute magnitude, thereby capturing an initially vigorous decline that gradually decelerates—this value marks the greatest deviation from the experimental reference. Conversely, the uniform reduction scenario (**Fig. 2b**) produces an L-Fit slope of *β*_1_ = −0.0126, which aligns more closely with the LS-Fit value and suggests a robust yet consistent clearance rate. Finally, the delayed-onset acceleration scenario (**Fig. 2c**) exhibits the shallowest L-Fit slope, *β*_1_ = −0.0104, indicative of a pronounced initial lag prior to the onset of accelerated decay. Collectively, these L-Fit *β* estimates—derived from the advanced GAN algorithm—underscore the distinct kinetic profiles across the scenarios while offering deeper quantitative insights beyond those provided by the constant experimental LS-Fit value.

## 3 Materials and Methods

### 3.1 Recombinant Protein Production

The S1 (2.1 kb) and S2 (2.3 kb) genes were PCR-amplified from plasmid VG40589-CY (Sino Biological Beijing, China) using primers flanked by BamHI/SalI restriction sites (in bold), see **Table S1**.

Digested products were ligated into pET28a (Novagen) and transformed into *E. coli* Rosetta(DE3) via heat shock (42°C, 45 s). Protein expression was induced with 1 mM isopropyl *β*-d-1-thiogalactopyranoside (IPTG) for 4 h at 37°C in LB medium containing 50 *μ*g/mL kanamycin and 35 *μ*g/mL chloramphenicol.

Cells were lysed in buffer containing 50 mM Tris-HCl (pH 8.0), 1 mg/mL lysozyme, 0.1% Triton X-100, and protease inhibitors (Roche). Inclusion bodies were solubilized in 8 M urea (pH 7.4) and purified by immobilized metal affinity chromatography (IMAC) using HisPrep FF 16/10 columns (Cytiva) on an ÄKTA Purifier system (GE Healthcare). Eluted fractions were analyzed by 12% SDS-PAGE and Western blotting with anti-His^®^ Tag monoclonal antibody (1:5,000; Sigma-Aldrich).

### 3.2 Expression of S1 and S2 Proteins in *E. coli* Cultures

The *Rossetta* strain of *E. coli* was transformed with the expression vectors `pET28a-S1` and `pET28a-S2` for the S2 protein. Bacterial cultures were grown under standard conditions in Luria-Bertani (LB) broth composed of 10 g/L tryptone, 10 g/L NaCl, and 5 g/L yeast extract, supplemented with 50 μg mL^−1^ kanamycin and 35 μg mL^−1^ chloramphenicol.

Recombinant protein expression was induced with 1 mM IPTG for 4 hours at 37 °C. Following induction, bacterial cells were collected by centrifugation and stored at −20 °C until further processing. The cell pellets obtained from centrifugation were resuspended in lysis buffer (50 mM Tris, *pH* 8, 0.5 M NaCl, 10 mM EDTA, 5 mM *β*-mercaptoethanol, and 0.35 mg/mL lysozyme) and incubated for 30 minutes at 30 °C. The suspension was then supplemented with 2% Triton X-100. Subsequently, 10 μg mL^−1^ RNase, 5 μg mL^−1^ DNase, and 6 mM MgCl_2_ were added, along with protease inhibitors and additional Triton X-100 to achieve a final concentration of 5%, and the mixture was incubated for 20 minutes.

Thereafter, the bacterial suspension was sonicated using an SLP^e^ Digital Sonifier (Branson) to ensure efficient cell disruption and release of inclusion bodies. The inclusion bodies were collected by centrifugation at 12 000 g for 30 minutes at 4 °C and subsequently washed under the same conditions with decreasing concentrations of Triton X-100 (2%, 1%, and 0.5%). Finally, three washes with PBS were performed before solubilizing the inclusion bodies in an 8 M urea buffer (50 mM phosphate buffer, *pH* 7, 0.3 M NaCl, and 8 M urea).

### 3.3 S1 Purification by Immobilized Metal Affinity Chromatography (IMAC)

IMAC chromatography was performed using an ÄKTA purification system (Cytiva, Marlborough, MA, USA) equipped with HisPrep FF 16/10 columns (Cytiva, Marlborough, MA, USA). The column was first equilibrated with 10 column volumes (CV) of binding buffer (50 mM phosphate buffer, pH 7, 0.3 M NaCl, 8 M urea). Solubilized inclusion bodies were clarified by centrifugation at 12 000 g for 30 minutes at 4 °C prior to loading onto the column.

Following sample application, the column was washed with 5 CV of binding buffer to remove unbound components. Elution was then performed stepwise using four sequential steps of 5 CV each with elution buffers containing 0%, 10%, 20%, and 100% of the elution buffer composition (50 mM phosphate buffer, pH 7, 0.3 M NaCl, 8 M urea, and 0.5 M imidazole). Eluted fractions were collected in 1 mL aliquots and subsequently concentrated via ultrafiltration using an Amicon Ultra-15 filter device with a molecular weight cutoff (MWCO) of 30 kDa (Millipore, Burlington, MA, USA).

The recombinant proteins obtained from this purification process were analyzed by SDS-PAGE. Additionally, Western blot analysis was performed using a mouse monoclonal anti-hexa-histidine (H6) antibody to confirm protein purification.

### 3.4 Polyclonal Antibody Production

Six-week-old male CD1 mice were immunized with purified recombinant proteins S1 and S2. The immunization regimen consisted of weekly injections for six consecutive weeks. The initial injection employed Freund’s complete adjuvant (Sigma-Aldrich, St. Louis, MO, USA), while the subsequent booster injections were administered with Freund’s incomplete adjuvant (Sigma-Aldrich, St. Louis, MO, USA). One week following the final immunization, mouse sera were collected and titrated via an indirect ELISA.

For the ELISA, microtiter plates were first incubated overnight at 4 °C under agitation to allow for antigen adsorption. After incubation, the plates were washed with PBS containing 0.1% Tween 20 to remove unbound antigens. The wells were then blocked with a solution of 2% skimmed milk in PBS-Tween 20 (0.1%) and agitated for 12 hours at 4 °C. Following blocking, the plates were washed at least three times with PBS-Tween 20. Thereafter, 100 μL of serial dilutions of the immunoserum, prepared in PBS, were added to the wells and incubated for 2 hours at room temperature under gentle agitation. Subsequent to further washing, the plates were incubated with a horseradish peroxidase-conjugated anti-mouse Ig antibody. Color development was achieved by adding a peroxidase substrate composed of 4 mg o-phenylenediamine dihydrochloride (OPD; Sigma-Aldrich, St. Louis, MO, USA) dissolved in 10 mL citrate-phosphate buffer (pH 5.0) with 4 μL of 30% H_2_O_2_ for 20 minutes. The reaction was stopped, and the absorbance was measured at 492 nm.

### 3.5 Automated Room Disinfection with DuctFIT^®^ Technology

The disinfection system employed in this study was based on the ductFIT^®^ technology developed by CleanAir Spaces. As illustrated in **Figure 3**, the hydrogen peroxide nebulization unit is strategically deployed within the room to ensure uniform disinfection. This system utilizes a patented photocatalytic process that combines catalytic components with a UV lamp operating at a specific frequency, resulting exclusively in the generation of hydrogen peroxide (H_2_O_2_) as the active disinfectant. Importantly, the system produces H_2_O_2_ at safe concentrations (0.03–0.05 ppm).

**Figure 3.**
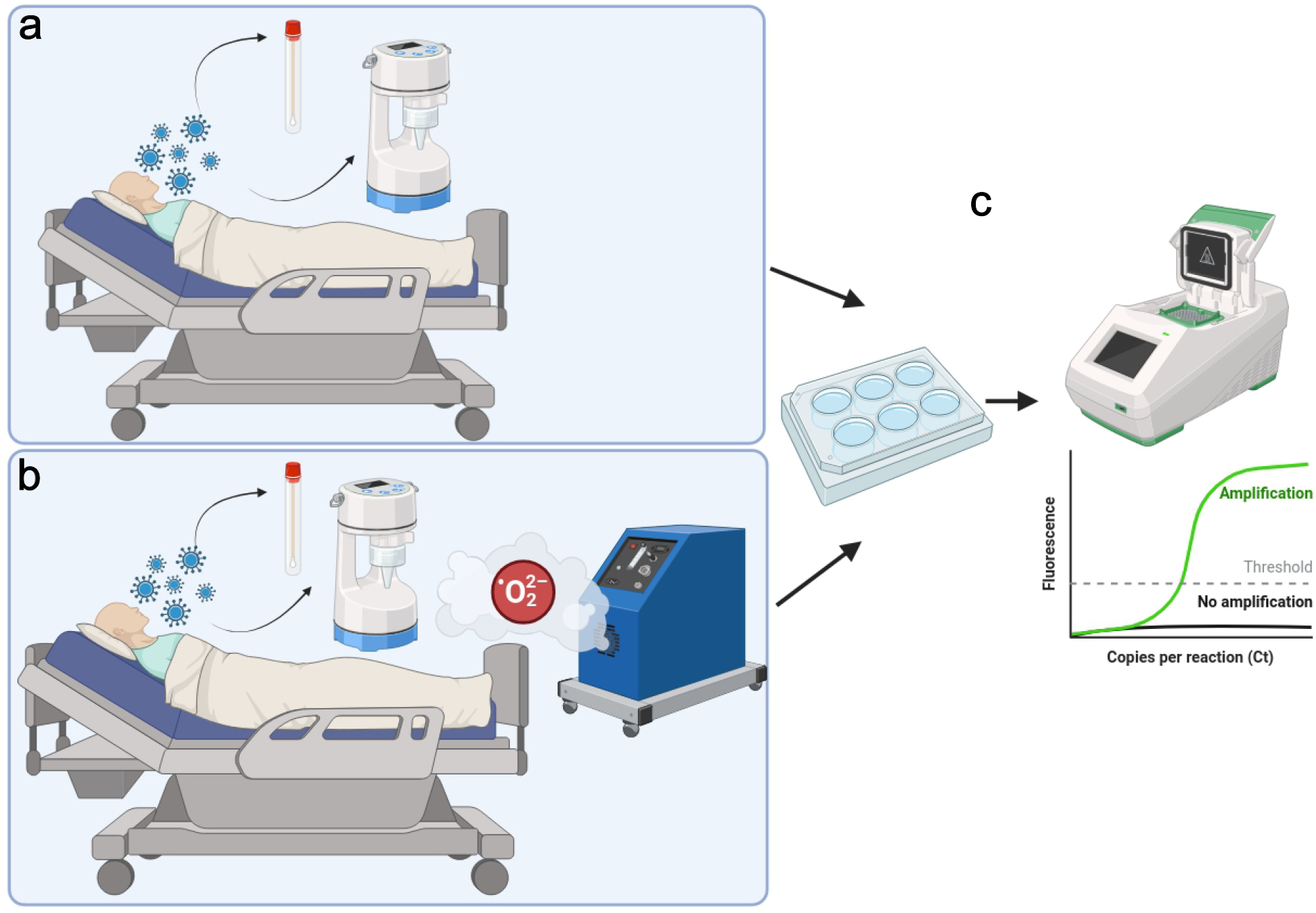
Schematic representation of the experimental workflow for evaluating automated hydrogen peroxide (H_2_O_2_) nebulization in a hospital setting. **(a)** Environmental sampling of air and surfaces in a patient room before decontamination, capturing SARS-CoV-2 particles for subsequent laboratory analysis. **(b)** Deployment of the H_2_O_2_ nebulization system, designed to reduce viral loads in the treated environment. **(c)** Assessment of residual infectivity and viral RNA levels via Vero E6 cell culture assays and RT-qPCR, with representative amplification curves illustrating threshold cycle (*C*_*t*_) values. This integrated approach provides both molecular and biological evidence of the decontamination protocol’s effectiveness. This illustration was created using Inkscape^29^.

For the clinical trial conducted in patient rooms, three ductFIT® Mobile units were strategically placed within a 90 m^2^ (252 m^3^) space. Operating at a medium airflow speed, each unit delivered a flow rate of 629 m^3^/h, resulting in a combined treatment capacity of 1887 m^3^/h and achieving 7.49 air changes per hour, thereby ensuring effective pathogen inactivation. Additionally, within the UCRI ward of Hospital Virgen de las Nieves, individual ductFIT® Mobile units were installed in rooms 115 and 116—areas lacking negative pressure—to evaluate the efficacy of H_2_O_2_ against SARS-CoV-2 under real-world hospital conditions.

### 3.6 Air and Surface Sampling Procedures for SARS-CoV-2 Assessment

Air and surface samples were collected from a hospital room before and after decontamination with an automated hydrogen peroxide (H_2_O_2_) nebulization system. Sampling was conducted by trained healthcare professionals under strict biosafety conditions to ensure reliable detection of SARS-CoV-2 contamination^28^.

Air samples were obtained using a Coriolis Compact Air Sampler (Bertin Technology) operating at a flow rate of 50 L/min for 20 minutes to capture airborne viral particles. Collected samples were immediately stored at 4 °C until further processing.

Surface samples were collected using sterile cotton swabs soaked in Hanks solution from a 25 cm^2^ area on surfaces including bed railings, tables, shelves, door handles, floors, and medical equipment. Following collection, the swabs were placed in Hank’s salt solution and stored at 4 °C until analysis. All samples were transported to a biosafety level 3 (BSL-3) laboratory, where RT-qPCR and viral culture assays were performed to evaluate both the presence and viability of SARS-CoV-2.

### 3.7 Biosafety Protocols

All SARS-CoV-2-related procedures were conducted within a Biosafety Level 3 (BSL-3) facility by highly trained personnel. These activities were executed in strict accordance with protocols approved by the institutional biosafety committee, thereby ensuring the safety of laboratory staff and preventing any environmental contamination^30^.

### 3.8 Virus Propagation and Sample Processing

Vero E6 cells (ECACC No. 84113001) were employed for viral propagation. Cells were maintained in Dulbecco’s Modified Eagle’s Medium (DMEM; Sigma-Aldrich, St. Louis, MO, USA) supplemented with 10% heat-inactivated fetal bovine serum (FBS; Gibco, Waltham, MA, USA)—which was inactivated at 56 °C for 30 minutes—along with penicillin (100 U/mL) and streptomycin (100 μg/mL) (Sigma-Aldrich, St. Louis, MO, USA). Cultures were incubated at 37 °C in a humidified atmosphere containing 5% CO_2_.

Air and surface samples were initially resuspended in 2 mL of Hank’s Balanced Salt Solution (HBSS) and then filtered through a 0.22 μm membrane filter (Sartorius, Göttingen, Germany) to remove potential contaminants, including bacteria and fungi. From the filtered samples, 500 μL aliquots were inoculated onto semi-confluent monolayers of Vero E6 cells—previously seeded at 5×10^5^ cells per well in a 6-well plate and incubated overnight—and further incubated for 1 hour under gentle agitation. Thereafter, 1 mL of Minimum Essential Medium (MEM) supplemented with 2% FBS was added, and the cultures were incubated at 37 °C for three days to facilitate viral replication.

Following the incubation period, the culture medium was aspirated, and 1 mL of TRIzol reagent (Thermo Fisher Scientific, Waltham, MA, USA) was added to each well. After a 15-minute incubation to ensure complete lysis, the TRIzol solution was transferred to RNase-free Eppendorf tubes and stored at −80 °C until further processing.

### 3.9 Indirect SARS-CoV-2 IgG ELISA

To detect SARS-CoV-2 infection, Vero E6 cells were seeded in 24-well plates and cultured in Dulbecco’s Modified Eagle’s Medium (DMEM; Sigma-Aldrich, St. Louis, MO, USA) supplemented with 10% heat-inactivated fetal bovine serum (FBS; Gibco, Waltham, MA, USA)—inactivated at 56 °C for 30 minutes—and antibiotics (penicillin at 100 U/mL and streptomycin at 100 μg/mL; Sigma-Aldrich, St. Louis, MO, USA). Cultures were maintained at 37 °C in a humidified atmosphere with 5% CO_2_.

After 24 hours, when infected cells exhibited cytopathological effects yet remained adherent, viral propagation was evaluated using a polyclonal anti-protein S2 serum. The culture medium was removed and cells were washed twice with sterile phosphate-buffered saline (PBS). Cells were then fixed with methanol at room temperature for 10 minutes and subsequently permeabilized by adding 100 μL of 0.1% NP40 in PBS for 1 hour at room temperature.

Following permeabilization, the plates were washed three times with PBS containing 0.5% Tween 20 (PBST) and then blocked overnight at 4 °C with a blocking buffer composed of PBS/0.1% Tween 20 and 2% milk powder. Mouse sera containing S2 polyclonal antibodies, diluted 1:500 in blocking buffer, were incubated with the cells for 2 hours at 37 °C. After four washes with PBST, a secondary anti-rat antibody conjugated with horseradish peroxidase (HRP) was applied at a 1:2000 dilution and incubated for 1 hour at 37 °C.

Subsequently, following four additional washes with PBST, a peroxidase substrate solution—comprising 4 mg of OPD dissolved in 10 mL of citrate-phosphate buffer (pH 5.0) with 4 μL of 30% H_2_O_2_—was added. After 5 minutes of incubation at room temperature, protected from light, the reaction was terminated by the addition of 0.5 M H_2_SO_4_. The optical density (OD) was measured at 450 nm with baseline correction at 620 nm using an Asys Expert 96 UV microplate reader (Thermo Scientific).

## Viral Recovery and RT-qPCR Analysis

Viral inactivation of SARS-CoV-2 on surfaces and environmental samples was assessed via RT-qPCR on propagated viral cultures in Vero E6 cells. Following viral propagation, RNA was extracted using the RNeasy Mini Kit (Qiagen, Hilden, Germany) in accordance with the manufacturer’s protocol.

Subsequently, a single-step multiplex real-time reverse transcription PCR (RT-qPCR) assay, employing fluorescence-labeled probes, was conducted using the Detection expert 1S SARS-CoV-2 kit (Genestore). Data acquisition was performed on a CFX Connect real-time PCR detection system (Bio-Rad) under the following cycling conditions: reverse transcription at 42 °C for 5 minutes, initial denaturation at 95 °C for 5 minutes, followed by 40 cycles of denaturation at 95 °C for 5 seconds and annealing at 58 °C for 15 seconds. The primer and probe sequences employed in this study are summarized in Table 2.

**Table 1.**
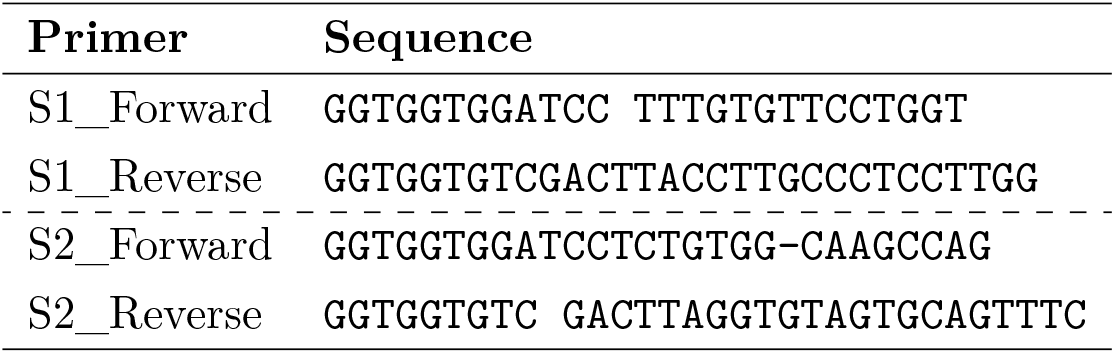
Primer sequences employed for the amplification of the S1 and S2 regions. These oligonucleotides were meticulously designed to ensure specificity and optimal performance during PCR amplification.

**Table 2.**
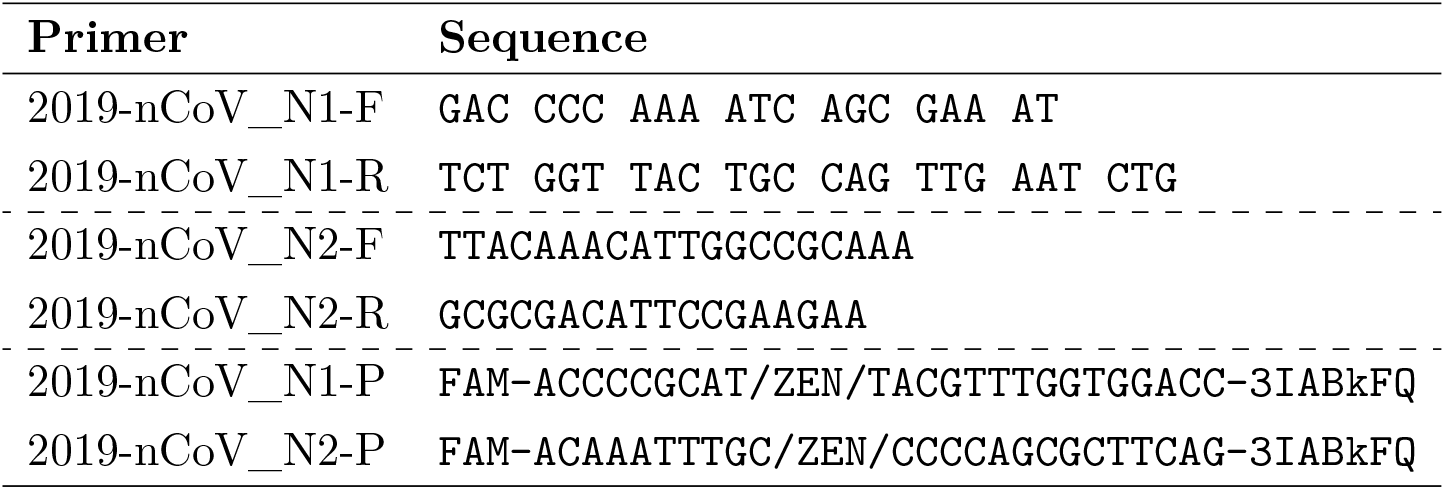
Primer sequences targeting the SARS-CoV-2 N gene regions (N1 and N2) used in RT-qPCR assays. The table includes both the forward and reverse primers as well as the corresponding probes, which were designed to maximize specificity and sensitivity for accurate viral detection.

### 3.10 Statistical and Modeling Analysis

The sample size (n = 72 total samples from 18 rooms) was determined through a priori power analysis based on a two-sample t-test design (*α* = 0.05, power = 80%) based on published SARS-CoV-2 environmental contamination data^31^. Assuming a mean difference in viral load (Δ*C*_t_ = 3–5) between pre- and post-decontamination conditions, as reported in similar studies^32,33,34^, our sample size was projected to detect a twofold difference in viral titers with 95% confidence. This aligns with the sample sizes used in comparable studies: Fu et al. (2012) examined 12 samples from a single hospital room^32^, Holmdahl et al. (2015) examined 96 samples from 4 rooms^33^, and Estienney et al. (2022) evaluated 32 samples from 32 rooms^34^.

Data are expressed as mean ± standard deviation (SD). Comparisons among multiple groups were conducted using one-way ANOVA followed by Tukey’s post-hoc test. For pairwise comparisons, an unpaired two-tailed Student’s t-test was applied after confirming normality with the Shapiro-Wilk test and homogeneity of variances with Levene’s test. Statistical significance was defined as *p <* 0.05. All analyses were performed using Python 3.9.

To characterize the temporal kinetics of H_2_O_2_ nebulization-mediated decontamination, a piecewise exponential model was employed, and parameters were estimated using non-linear least squares fitting. Furthermore, a Generative Adversarial Network (GAN) was utilized to generate synthetic data points, thereby validating the robustness and predictive capacity of our model.

## 4 Discussion

The results of this study demonstrate the significant efficacy of hydrogen peroxide (H_2_O_2_) nebulization in reducing SARS-CoV-2 contamination in hospital environments. Prior to decontamination, 55.6% of air samples and 44.4% of surface swabs tested positive for viral RNA, with cycle threshold (C_*t*_) values indicative of moderate to high viral loads. These findings are in close agreement with previous reports of widespread environmental contamination in COVID-19 patient rooms. For instance, Jones et al.^35^ detected SARS-CoV-2 RNA in 56.7% of air samples and 47.5% of surface swabs, while Andersen et al.^36^ reported comparable contamination rates in intensive care units. Moreover, the virological assessment by Wolfel et al.^1^ and the aerosol and surface stability study by van Doremalen et al.^2^ substantiate the persistence of SARS-CoV-2 in clinical settings. In addition, Chin et al.^3^ demonstrated the virus’s robustness under varying environmental conditions, thereby reinforcing the critical need for effective decontamination protocols. The established principles of antisepsis and disinfection^4,9^ further underscore that robust H_2_O_2_ treatments can mitigate environmental viral loads, as observed in our study. **Figure 3** succinctly summarizes the methodological workflow, providing context for our discussion on the effectiveness of the automated decontamination system and its impact on viral inactivation.

Following H_2_O_2_ nebulization, our data reveal a statistically significant reduction in viral RNA detection. Post-treatment, only 22.2% of air samples and 13.9% of surface swabs remained positive, with markedly increased C_*t*_ values indicative of substantially diminished viral loads. This outcome aligns with the findings of Zhang et al.^34^, who reported a 3–5 unit increase in C_*t*_ values after H_2_O_2_ treatment—corresponding to an approximate 90% reduction in viral RNA. Additionally, studies by López Ortega et al.^10^ and Goyal et al.^11^ have demonstrated the virucidal efficacy of hydrogen peroxide in various formulations, while Fadaei^15^ confirmed that dry-mist disinfection with H_2_O_2_ effectively inactivates human coronaviruses. These results are also in concordance with broader decontamination reviews^14,13^ and practical assessments of hydrogen peroxide vapor systems^8,5**?**, 6,7^.

Complementary to the molecular data, our investigation into viral infectivity—evaluated through Vero E6 cell cultures and an indirect ELISA targeting the SARS-CoV-2 S2 protein—further substantiates the treatment’s efficacy. Prior to decontamination, 34.7% of the cultures exhibited cytopathic effects (CPE), correlating strongly with low C_*t*_ values (i.e., *<*38) and the presence of infectious virus, as reported by Wolfel et al.^1^ and Bullard et al.^20^. Following H_2_O_2_ nebulization, only 5.6% of cultures demonstrated minimal CPE, accompanied by an approximately threefold reduction in ELISA optical density values. This dramatic decline in both infectious virus and viral antigen levels suggests that the residual viral RNA detected at high C_*t*_ values likely represents non-viable viral particles^37^.

Interestingly, our findings indicate that environmental contamination was not universal among patients, likely due to variability in individual viral excretion dynamics and environmental factors such as airflow, surface characteristics, and patient activity levels^38^. Nonetheless, once contamination occurred, the deployment of automated H_2_O_2_ nebulization proved highly effective in dramatically reducing infectivity, as demonstrated by both RT-qPCR and ELISA assays. The significant decrease in the proportion of positive samples, accompanied by increased C_*t*_ values, underscores a marked reduction in viral load, while the pronounced decline in viral antigen levels further validates the elimination of infective particles.

Real-time reverse transcription polymerase chain reaction (RT-qPCR), executed according to protocols developed by the National Institute of Infectious Diseases (NIID) in Japan, reliably detected SARS-CoV-2 RNA at two nucleocapsid (N) gene target sites with high sensitivity and specificity^39^. Furthermore, the utilization of Vero E6 cell cultures allowed us to discern only those viral particles capable of infection, ensuring that our infectivity assays reflect viable virus. Enzyme-linked immunosorbent assays targeting the S2 protein provided complementary evidence, confirming the presence of viral antigens and the capacity of the virus to multiply in these cells^40^. Collectively, this integrated approach is well-suited for quantifying SARS-CoV-2 from both air and surface samples in hospital settings, yielding valuable insights into environmental viral loads and informing infection control strategies^41^.

SARS-CoV-2 RNA was quantified by RT-qPCR in only 24 of the 72 samples, suggesting that the viral load present in individual patients directly influences environmental contamination. Previous studies have shown that patients with high viral loads or overt respiratory symptoms (e.g., coughing, sneezing) are more likely to disseminate infectious particles through the air and onto surfaces^35,42,43,36,34^. Conversely, asymptomatic patients or those with low viral loads may contribute minimally to environmental contamination^44^. Notably, our findings indicate that once environmental contamination occurs, automated H_2_O_2_ nebulization is highly effective in drastically reducing both viral RNA and antigen levels. The observed reduction in positive sample rates and the elevation in C_*t*_ values, along with the significant drop in ELISA-detected viral proteins, collectively underscore the ability of H_2_O_2_ nebulization to diminish the infectivity of SARS-CoV-2 in clinical environments.

Extrapolating from these empirically derived kinetic models to real-world decontamination practice reveals significant implications for protocol design and implementation. The temporal scale delineated in **Figure 2** (0–48 hours) strategically aligns with critical operational phases within standard environmental decontamination protocols, encompassing pre-treatment, active disinfection, and post-treatment validation. In the *rapid-onset with progressive deceleration* scenario, the moderate delay time (*τ* ~ 7.8 h) suggests that although the virucidal effect of H_2_O_2_ initiates relatively quickly, a finite interval is required to achieve the most pronounced reduction in viral load. By contrast, the *delayed onset with accelerated decay* scenario exhibits a substantially longer delay period (*τ* ~ 25.9 h) but a more vigorous eventual decline, underscoring the potential for environmental or procedural factors—such as localized humidity or airflow patterns—to postpone the onset of effective H_2_O_2_ action before a steep reduction phase ensues. Meanwhile, the *uniform reduction* scenario, characterized by *τ* ~ 10.0 h, reflects a nearly constant rate of viral decay once nebulization begins, thereby highlighting conditions in which neither early saturation nor delayed activation dominates the disinfection kinetics.

These results align well with foundational studies by van Doremalen *et al*.^2^ and Chin *et al*.^3^, who demonstrate that natural aerosol sedimentation, ventilation strategies, and ambient factors can influence viral decay rates in indoor settings. Moreover, Kampf^12^ and Otter *et al*.^45^ have emphasized that humidity, temperature, and airflow patterns interact with disinfectant aerosols, potentially modulating the effective onset time of virucidal action. In enclosed or high-humidity environments (where concave kinetics predominate), a lengthier nebulization period or higher H_2_O_2_ concentration may be required to overcome initial inhibitory effects and achieve meaningful viral load reduction. Conversely, in well-ventilated spaces conducive to rapid mixing and dispersion (conditions favoring convex or linear kinetics), shorter nebulization cycles could suffice, thereby optimizing resource use without compromising disinfection efficacy.

Taken together, these piecewise exponential models furnish a robust, data-driven framework for tailoring H_2_O_2_ nebulization protocols to diverse clinical environments. The inclusion of both a rate constant (*k*) and a delay time (*τ*) in the modeling scheme enables a nuanced understanding of how decontamination unfolds across different temporal segments, thereby informing evidence-based modifications to exposure duration, aerosol density, and operational logistics. In this regard, the superior performance of the non-linear fits—consistently higher *R*^2^ values relative to linear approximations—underscores the importance of incorporating non-linear dynamics in future disinfection studies. Ultimately, these findings reinforce the centrality of context-specific strategies, supporting a paradigm in which H_2_O_2_ nebulization cycles are judiciously matched to local architectural and environmental conditions to maximize viral inactivation while minimizing unnecessary downtime or chemical usage^46^.

The incorporation of Generative Adversarial Networks (GANs) into viral decay simulations, as depicted in **Figure 2**, represents a significant leap forward in data augmentation for kinetic modeling. By leveraging advanced machine learning algorithms, the synthetic data produced by GANs—overlaid onto experimental measurements—bridges critical gaps and fortifies the robustness of our analyses. This sophisticated approach is particularly advantageous in instances where empirical observations are limited or constrained by experimental challenges, as GAN-derived datasets provide a richer, more nuanced perspective on viral decay dynamics. The heightened granularity conferred by these synthetic data not only bolsters the statistical rigor of the findings but also enables a more thorough validation of proposed kinetic models. In doing so, the integration of intelligent algorithms underscores a promising trajectory in virological research, offering deeper insights into complex biological processes and elevating the overall precision of kinetic modeling.

## 5 Limitations

While this study provides valuable insights into the efficacy of H_2_O_2_ nebulization and the utility of recombinant SARS-CoV-2 subunits, several limitations should be acknowledged. First, the sample size, though sufficient for detecting significant differences, may not capture all sources of variability in environmental contamination across diverse clinical settings; future studies should include a larger and more diverse set of sampling locations to enhance generalizability. Second, the use of *E. coli* for protein expression, while advantageous for yield and scalability, limits our ability to produce glycosylated proteins, which may be critical for fully recapitulating native antigenic structures. Future work should explore eukaryotic expression systems to overcome this limitation and provide proteins with post-translational modifications. Finally, the clinical significance of residual viral RNA post-decontamination remains uncertain, warranting further investigation to determine whether these low-level signals represent a meaningful transmission risk, which could be addressed by incorporating viability assays and correlating molecular findings with infectivity studies.

## 6 Conclusions and Future Directions

Our findings demonstrate that H_2_O_2_ nebulization effectively reduces SARS-CoV-2 contamination, as evidenced by significant decreases in both viral RNA and antigen levels. The application of a piecewise exponential model—integrating a delay parameter and curvature factor—provides a robust quantitative framework for characterizing the non-linear kinetics of decontamination. These results align with seminal studies^2,3,12^ and underscore the potential for tailoring disinfection protocols based on environmental and operational variables.

Future studies should explore eukaryotic expression systems to overcome the limitations associated with non-glycosylated protein production in *E. coli*, thereby enhancing the fidelity of recombinant antigens for diagnostic and immunological applications. Moreover, further investigation into the clinical relevance of residual viral RNA is needed to ascertain whether these trace levels pose a genuine risk for transmission, ultimately guiding the refinement and personalization of H_2_O_2_ nebulization protocols in healthcare environments.

Unlike previous studies where sample collection was typically centralized (Wölfel et al., 2020^1^; Zou et al., 2020^47^), this investigation uniquely conducted sampling with patients remaining in their own hospital rooms. This novel approach minimizes environmental variability and better captures the authentic in situ viral dynamics, thereby offering critical insights into viral behavior and transmission in a clinical context. The findings underscore the value of context-specific sampling strategies and have significant implications for refining infection control practices.

Notably, our study employs an advanced Generative Adversarial Network (GAN) algorithm—a methodology scarcely applied in the realm of viral kinetics^27,48^—to generate synthetic data that substantially bolsters the validation and robustness of our statistical models. This intelligent algorithmic approach not only enriches the dataset but also enhances parameter estimation and model verification, thereby mitigating the limitations imposed by sparse experimental data. In leveraging GAN-derived synthetic data, we pave a promising path forward in virological research, offering refined experimental designs and deeper insights into complex viral decay dynamics, ultimately advancing our collective understanding of these critical processes.

## Supporting information

SupportingInformation_SHAKEN

## Data Availability

All datasets generated and/or analysed during the current study are publicly accessible. These data are hosted on the GitHub platform at https://github.com/renee29/pyH2O2DeconNeb_KineticsModeling.git. For long-term archival and citation, the data are also deposited in Zenodo under the digital object identifier https://doi.org/10.5281/zenodo.15536388.

https://doi.org/10.5281/zenodo.15536388

## 8 Ethical Approval

Ethical approval for this study, designated internally as Protocol ProjectDuctFit-V3 (Version V.3), was granted by the CEIM/CEI Provincial de Granada (Comité de Ética de la Investigación con Medicamentos / Comité de Ética de la Investigación Provincial de Granada) on July 20, 2020 (Act No. 8/20), with the official certificate dated September 23, 2020.

## 10 Acknowledgments

This paper has been partially supported by Grant PID2022-137228OB-I00, funded by the Spanish Ministerio de Ciencia, Innovación y Universidades (MICIU/AEI/10.13039/501100011033 & “ERDF/EU A way of making Europe”); by Grant C-EXP-265-UGR23, funded by the Consejería de Universidad, Investigación e Innovación & ERDF/EU Andalusia Program; by the Modeling Nature Research Unit, project QUAL21-011; and by the María Zambrano-Senior grant, provided by the Spanish Ministerio de Universidades and Next-Generation EU.

## Code Availability

The Python code developed for the kinetics study is publicly available in the following SciHub repository:

https://github.com/renee29/pyH2O2DeconNeb_KineticsModeling.git

Researchers are encouraged to explore and adapt this code for their own analyses, as it provides a comprehensive framework for modeling and interpreting kinetic data in related fields. Should you have any inquiries or require additional assistance, please contact the corresponding authors directly.

## Author Contributions

C.M.-G. and R.F. conceived and designed the research. J.A.S.-M., B.C.G.-H. and P.G.-V. coordinated sample collection and managed patient logistics and facility operations at the hospital. J.S.-P., M.B., A.F., A.C.-G. and A.O.-C.-A. processed viral RNA using RT-qPCR, quantified viral antigens via indirect ELISA, and assessed infectivity through Vero E6 cell cultures. R.F. conducted the statistical analyses, developed and implemented the kinetic modeling and simulations. R.F., J.S.-P., C.M.-G., J.A.S.-M., and A.O.-C.-A. refined the study design and wrote the manuscript. All authors participated in discussions, provided critical revisions, and approved the final version of the manuscript.

