## SupportingInformation_SHAKEN for "Exploring Novel Kinetics of Automated H_2_O_2_ Nebulization: A Breakthrough in SARS-CoV-2 Elimination"

Tuesday 3rd June, 2025

### 1 ELISA OD Measurements versus Control.

Table S1 presents representative ELISA results, reporting optical density (OD) values relative to a control and their corresponding standard deviations (SD). In our study, air samples from patients were assayed both before and after $H_2O_2$ nebulization. Notably, Patient 2's pre-treatment air sample exhibited a high OD of 0.7443 ± 0.3734, which decreased to 0.3196 ± 0.0428 following treatment—consistent with RT-qPCR data indicating a reduction in viral load. Patient 1, while showing a more modest decline (from 0.1639 ± 0.3750 to 0.1420 ± 0.2698), similarly demonstrated reduced antigen presence post-treatment. These findings substantiate the efficacy of $H_2O_2$ nebulization and align with previous reports[1,2].

| **Sample** | **OD vs. Control ± SD** |
|---|---|
| Patient 1 – Air (No $H_2O_2$) | 0.1639 ± 0.3750 |
| Patient 1 – Air (With $H_2O_2$) | 0.1420 ± 0.2698 |
| Patient 2 – Air (No $H_2O_2$) | 0.7443 ± 0.3734 |
| Patient 2 – Air (With $H_2O_2$) | 0.3196 ± 0.0428 |

**Table S1|** Representative ELISA OD measurements versus control and corresponding standard deviations (SD) from S2-based assays, conducted before and after $H_2O_2$ nebulization.

### 2 Duplicate ELISA Measurements.

Table S2 presents a subset of duplicate ELISA measurements of optical density (OD) values, recorded as "Duplicate 1" and "Duplicate 2", for samples collected from patients both before (bt) and after (at) $H_2O_2$ nebulization. These replicate measurements are crucial for assessing the intra-assay precision and reproducibility of the ELISA method used to quantify SARS-CoV-2 antigen levels. For example, Patient 1's untreated air sample yielded OD values of 1.4277 and 0.8973, while after treatment, the values decreased to 1.3314 and 0.9499. Similarly, Patient 2's air sample prior to treatment exhibited an OD of 2.0070 ± 1.4789, which decreased to 1.3485 ± 1.2879 following treatment—an observation that aligns with the documented reduction in viral infectivity. An additional "Extra case" is included as a reference control. Such reproducibility in the OD readings supports the reliability of our assay,

‡ These authors contributed equally.

consistent with the methodologies reported in the literature[3,4]. A detailed statistical summary is provided in Table S2.

| Sample | Duplicate 1 | Duplicate 2 |
|---|---|---|
| Patient 1 – Air (No $H_2O_2$) | 1.4276796 | 0.8973299 |
| Patient 1 – Air (With $H_2O_2$) | 1.3314332 | 0.9498714 |
| Patient 2 – Air (No $H_2O_2$) | 2.0070044 | 1.4788834 |
| Patient 2 – Air (With $H_2O_2$) | 1.3485021 | 1.2879408 |
| (Extra case) | 1.017415211 | 0.979823574 |

**Table S2| Duplicate ELISA Measurements for Air Samples.** This table presents replicate optical density (OD) readings for air samples collected from patients before and after $H_2O_2$ nebulization.

### 3 Cycle Threshold ($C_t$) values for Patients P1–P6.

Table S3 presents a subset of Cycle Threshold ($C_t$) measurements (reported as mean ± standard deviation (SD)) for six patients (P1–P6) under various sampling conditions. In the table, "A" denotes air samples and "S" denotes surface swabs, with "bt" indicating samples collected before treatment and "at" after $H_2O_2$ nebulization. Notably, several conditions exhibit undetectable viral RNA (recorded as 0), particularly post-treatment, which is consistent with a marked reduction in viral load.

For example, patient P1's data show a $C_t$ value of 25.00 ± 1.41 in the air sample before treatment, which increases to 29.42 ± 0.68 after treatment, while surface samples remain undetectable. Similarly, patient P3 demonstrates a $C_t$ of 29.82 ± 1.45 for surface swabs before treatment that increases to 34.15 ± 4.53 after treatment. These findings underscore the efficacy of $H_2O_2$ nebulization in reducing detectable viral RNA, in agreement with our broader observations.

| Patient | Condition | $C_t$ ± SD |
|---|---|---|
| P1 | Abt | 25.00 ± 1.41 |
| P1 | Aat | 29.42 ± 0.68 |
| P1 | Sbt | 0.00 ± 0.00 |
| P1 | Sat | 0.00 ± 0.00 |
| P2 | Abt | 23.48 ± 0.68 |
| P2 | Aat | 0.00 ± 0.00 |
| P2 | Sbt | 31.87 ± 1.41 |
| P2 | Sat | 0.00 ± 0.00 |
| P3 | Abt | 25.02 ± 0.08 |
| P3 | Aat | 26.32 ± 2.10 |
| P3 | Sbt | 29.82 ± 1.45 |
| P3 | Sat | 34.15 ± 4.53 |

| Patient | Condition | $C_t$ ± SD |
|---|---|---|
| P4 | Abt | 26.31 ± 0.92 |
| P4 | Aat | 29.93 ± 0.09 |
| P4 | Sbt | 33.46 ± 3.53 |
| P4 | Sat | 34.90 ± 2.79 |
| P5 | Abt | 36.39 ± 3.40 |
| P5 | Aat | 0.00 ± 0.00 |
| P5 | Sbt | 32.28 ± 0.77 |
| P5 | Sat | 0.00 ± 0.00 |
| P6 | Abt | 31.38 ± 0.81 |
| P6 | Aat | 31.91 ± 1.53 |
| P6 | Sbt | 24.79 ± 0.16 |
| P6 | Sat | 25.41 ± 0.59 |

**Table S3| Ct Measurements for Patients P1–P6.** Values are reported as: $C_t$ ± SD. Abbreviations: A = air, S = surface, bt = before treatment, and at = after $H_2O_2$ nebulization. Undetectable signals are recorded as 0.

### Mathematical Modeling of $H_2O_2$ Nebulization Kinetics.

In order to quantify the kinetics of $H_2O_2$ nebulization in reducing SARS-CoV-2 viral loads, we adopted a piecewise exponential model that effectively captures the two-phase decontamination process observed in our experiments. The model is defined as:

$$N(t) = \begin{cases} N_0, & \text{if } t < \tau, \\ N_0\, e^{-k(t-\tau)}, & \text{if } t \geq \tau, \end{cases}$$

where $N(t)$ represents the viral load at time $t$, $N_0$ is the initial viral load, $k$ is the rate constant determining the exponential decay, and $\tau$ is the delay time before the onset of decontamination.

The decontamination phase for $t \geq \tau$ is governed by the following ordinary differential equation (ODE):

$$\frac{dN(t)}{dt} = -k\,N(t),$$

with the initial condition $N(\tau) = N_0$. This classic model, which assumes that the viral decay rate is proportional to the current viral load[5,6], can be solved using standard techniques found in textbooks such as Boyce and DiPrima[7] and Coddington and Levinson[8]. Without detailing the integration steps, the solution for $t \geq \tau$ is given by

$$N(t) = N_0\,e^{-k(t-\tau)}.$$

For $t < \tau$, the model assumes that the viral load remains constant at $N_0$, reflecting the initial delay before the decontamination effect takes hold. This modeling approach provides a robust framework for capturing the kinetics of $H_2O_2$ decontamination, in line with empirical findings reported by van Doremalen et al.[1] and Chin et al.[2].

This piecewise exponential model is particularly useful because it accounts for the initial period during which the viral concentration remains unchanged—possibly due to factors such as natural aerosol settling or delayed activation of the nebulization process—and then captures the rapid exponential decay once $H_2O_2$ nebulization becomes effective. The applicability of this modeling approach has been supported by previous work; for instance, van Doremalen et al.[1] and Chin et al.[?] demonstrated that exponential decay models are suitable for evaluating viral stability on surfaces and in aerosols. Foundational texts by McDonnell[5] and Block[6] provide the theoretical basis for these kinetics in disinfection protocols, while more recent studies by Viana Martins et al.[9] and Cimolai[10] highlight the utility of such models for optimizing decontamination strategies. Collectively, these findings validate our piecewise exponential model as a robust and predictive tool for tailoring $H_2O_2$ nebulization strategies in healthcare settings.
